# Injection Molded Autoclavable, Scalable, Conformable (iMASC) system for aerosol-based protection

**DOI:** 10.1101/2020.04.03.20052688

**Authors:** James D. Byrne, Adam J. Wentworth, Peter R. Chai, Hen-Wei Huang, Sahab Babaee, Canchen Li, Sarah L. Becker, Caitlynn Tov, Soekkee Min, Giovanni Traverso

**Affiliations:** Harvard Radiation Oncology Program, Brigham and Women’s Hospital, Harvard Medical School, Boston, MA 02114, USA; Division of Gastroenterology, Brigham and Women’s Hospital, Harvard Medical School, Boston, MA 02115, USA; David H. Koch Institute for Integrative Cancer Research, Massachusetts Institute of Technology, Cambridge, MA 02142, USA; Division of Medical Toxicology, Department of Emergency Medicine, Brigham and Women’s Hospital, Harvard Medical School, Boston, MA; The Fenway Institute, Boston MA; Department of Psychosocial Oncology and Palliative Care, Dana Farber Cancer Institute, Boston MA; Department of Mechanical Engineering, Massachusetts Institute of Technology, 77 Massachusetts Ave, Cambridge, MA 02139, USA

## Abstract

There is a dire need for personal protective equipment (PPE) within healthcare settings during the COVID-19 pandemic. In particular, single use disposable N95 face masks have been limited in supply. We have developed an Injection Molded Autoclavable, Scalable, Conformable (iMASC) system for aerosol-based protection. The iMASC system was designed as a reusable liquid silicone rubber mask with disposable N95 filter cartridges that can fit most face sizes and shapes. This system reduced the amount of N95 filter while preserving breathability and fit. Using finite element analysis, we demonstrated mask deformation and reaction forces from facial scans of twenty different wearers. In addition, we validated these findings by successful fit testing in twenty participants in a prospective clinical trial. The iMASC system has the potential to protect our healthcare workers with a reusable N95-comparable face mask that is rapidly scalable.

## Introduction

Dwindling supplies of personal protective equipment (PPE) in hospitals is forcing healthcare workers to reuse and clean PPE using anecdotal strategies, which may weaken the effectiveness of PPE in protecting workers from acquisition of COVID-19 disease. In some places, the complete lack of PPE has resulted in healthcare workers using PPE that may have variable droplet protection (*1*). Shortages of PPE have significant impact among healthcare workers who evaluate individuals with suspected and confirmed COVID-19 disease (*1-2*). First, individuals using PPE acquired outside of the hospital may inadvertently be using PPE without droplet protection resulting in inadequate protection. Second, workers without PPE will acquire infections, including COVID-19, at greater rates than those with adequate PPE (*3*). Infected healthcare workers may transmit disease to family members, worsening the pandemic (*4*). Third, with increased COVID-19 infection among healthcare workers, the available workforce to address sick patients decreases, resulting in increasing morbidity and mortality (*4*). There is therefore a critical need to develop innovative measures to generate safe, reusable PPE.

Thus, we have designed and fabricated an Injection Molded Autoclavable, Scalable, Conformable (iMASC) system for aerosol-based protection with N95 material filters that can be inserted and replaced as needed. To understand the ability of our mask to conform to multiple face sizes and shapes, we have undertaken finite element analysis evaluating the deformability of the iMASC system. Lastly, we performed a prospective clinical trial for fit testing of our mask as well as qualitative assessment of the mask compared to the current N95 masks. Our goal is to address the critical shortage of N95 face masks to maximally protect healthcare workers and provide an enduring supply chain of N95 face masks to reduce and prevent COVID-19 transmission among healthcare workers and patients.

## Results

### Design and generation of injection molded liquid silicone rubber mask

The iMASC system was designed to function as an N95-comparable mask (**Fig. 1**). The shape of the iMASC system was modeled from disposable regular N95 masks used in the hospital, which are amenable to many different face sizes and shapes. Medical grade liquid silicone rubber (LSR) was identified as an optimal material for mask fabrication due to its conformable capacity, sterilizability through multiple methods and compatibility with injection molding for fabrication scalability. The weight of the iMASC system was 44.84 ± 0.05 grams (n = 3) compared to 10.41 ± 0.13 grams (n = 3) of current N95 masks. We employed a dual filter approach similar to half-mask elastomeric respirators to increase breathability and filtration area (*5*). A single regular N95 mask generated up to 5 filters for the iMASC system, thus extending the N95 material use. Furthermore, based upon the material selection of a medical grade LSR, the iMASC system is reusable after sterilization by cleaning with hospital grade bleach/alcohol wipes, autoclave and heating methods.

**Fig. 1.**
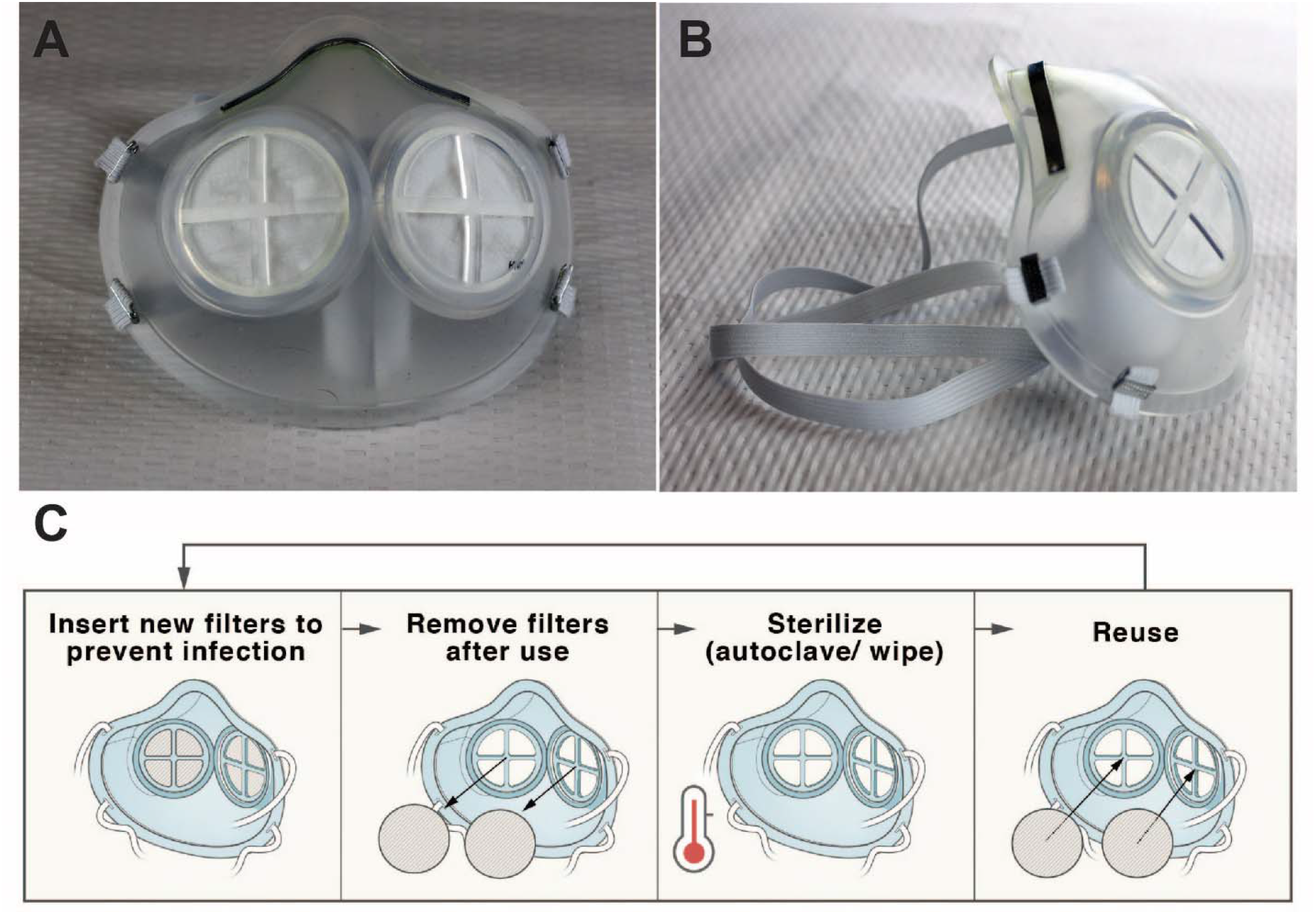
iMASC system for aerosol-based protection. (A) Front and (B) side images of the iMASC system. (C) Workflow for sterilization and reuse of iMASC system.

### Characterization of mask material after sterilization

An advantage of the iMASC system over the half-mask respirators is the methods of sterilization (see **table S1**). We have performed tensile tests of the mask material after 10 autoclave cycles and 5 minutes in a 1:10 bleach solution and 70% isopropyl alcohol. We found that 10 autoclave cycles make the mask slightly stiffer, while the bleach soak resulted in no change and the isopropanol alcohol soak makes the material less stiff (**fig. S1**). Despite these small changes in tensile strength, there were no gross differences in the mask compared to the non-sterilized mask.

### Finite element analysis for mask deformation upon different face shapes and sizes

We used non□linear finite element (FE) analyses (see “Deformation studies” in Methods) to evaluate the deformation response of the flexible mask frames while wearing and determine the forces required to keep the mask in place across a range of subject faces. In **Fig. 2A**, we reported the numerical snapshots of the face mask when subjected to the strap’s tensile loads, denoted by *T* shown in **fig. S3**, and monitored the deformation of the mask at different levels of the reaction force exerted from the mask to the face, *F =* 0 (undeformed), 4.5 (initial contact), and 10 (full contact) N. The color maps represent the distribution of displacement’s magnitude, *U*, showing relatively large deformation of the mask required to fit in to the subject face. We also calculated the normal contact forces, *F*^*N*^, and contact pressures, *P*, as a function of *F* to evaluate the interaction between the mask and face. In **Fig. 2B**, the distribution of the *F*^*N*^ are shown at the different *F*. As expected, no *F*^*N*^ was recorded <u>a</u>t *F =* 0. By pulling the straps, the mask starts to be engaged with the face, and at *F =* 4.5 N the maximum *F*^*N*^ occurs around the cheek. Further pulling the straps (*F =* 10 N) induces a relatively higher *F*^*N*^ along the edge of the mask in the check and chin (lower lips) rather than the nose and cheekbones. This is a signature of the need to the Aluminum strip to bond across the bridge of the nose to enhance the contact pressure.

**Fig. 2.**
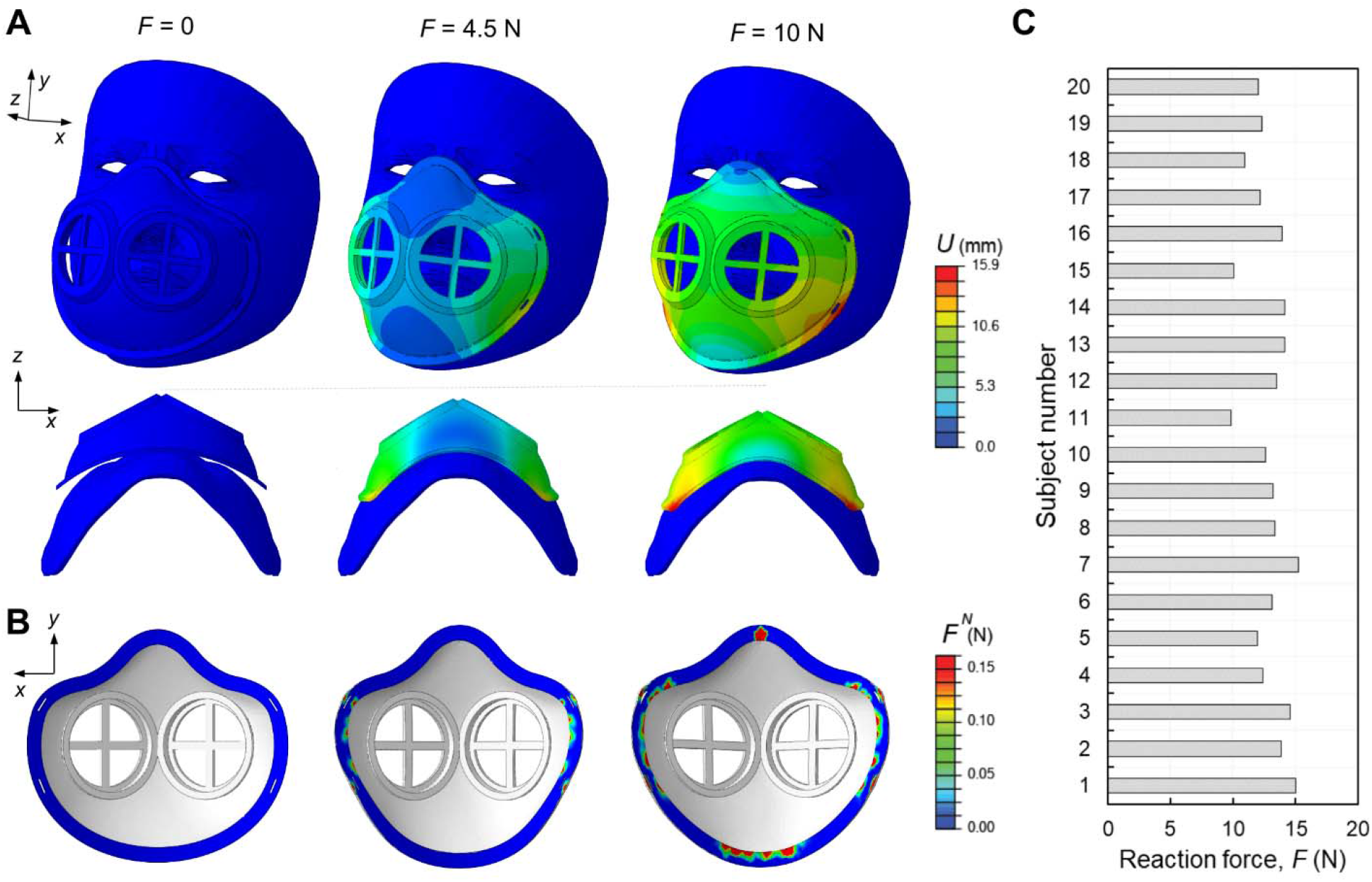
Finite Element modeling of flexible masks. (A) Numerical images showing the deformation of the elastomeric mask at different levels of reaction forces, F= 0, 4.5, and 10 N in two different views (top and bottom rows). The colors represent the magnitude of displacement field, U. (B) The corresponding distribution of the normal contact forces, F^N, between the mask and face. (C) Reaction forces for the subject numbers n=1,2,3,.., 20 computed from simulations.

Next, we estimated the reaction force required to achieve an average contact pressure of *P =*10 KPa (relatively uniformly distributed along the edge of the mask) as a higher limit of the contact pressure that results in a suitable fit between the mask and skin faces (*6*). This reaction force is equivalent to the force applied through the straps. In **Fig. 2C**, we reported the reaction forces for twenty different subjects, ranging from 9.5 to 15 N. These variations are duo to the difference in shape and size of the subject’s faces especially in the jaw and cheekbone parts. Through application of these forces via the straps combined with the aluminum strip across the nose bridge, one can guarantee the mask will be tightly stayed in place.

### Clinical trial evaluating mask fitting

In a prospective trial, we enrolled 24 healthcare workers at a large, urban, academic medical center who had been previously certified to wear a N95 respirator into our IRB-approved study. We excluded individuals with significant facial hair or those that had failed a N95 fit test. Consenting individuals were subject to a fit test as defined by the United States Occupational Safety and Health Administration (OSHA) (*7-8*). Briefly, participants first placed the iMASC system on their face and molded the nosepiece to ensure an adequate seal. Next, the participant’s head and face were placed in a plastic hood, and a saccharine solution was sprayed into the enclosed space as guided by OSHA (*7*). Participants were asked to perform four maneuvers: 1) rotating head in the lateral plane, 2) moving the head up and down, 3) verbally counting down backwards from 100 to 90 and 4) bending at the waist. A passing test was defined as no detection of saccharine solution by study participants. **Fig. 3A** shows the demographics of the participants, and **figs. S2** and **S3** showcase the 3D facial reconstructions demonstrating variability of facial sizes and shapes among the participants. The average age of participants was 41 years with a range of 21-65 years with an average BMI of 26.5. The breakdown of participants by profession was 46% nurses (n=11), 21% attending physicians (n=5), 21% resident physicians (n=5), and 12% technicians (n=3). Of these participants, 4 did not perform the fit testing (1 due to inability to detect saccharin solution on pre-mask placement sensitivity test, 2 due to time, and 1 due to fit of the mask on her face).

**Fig. 3.**
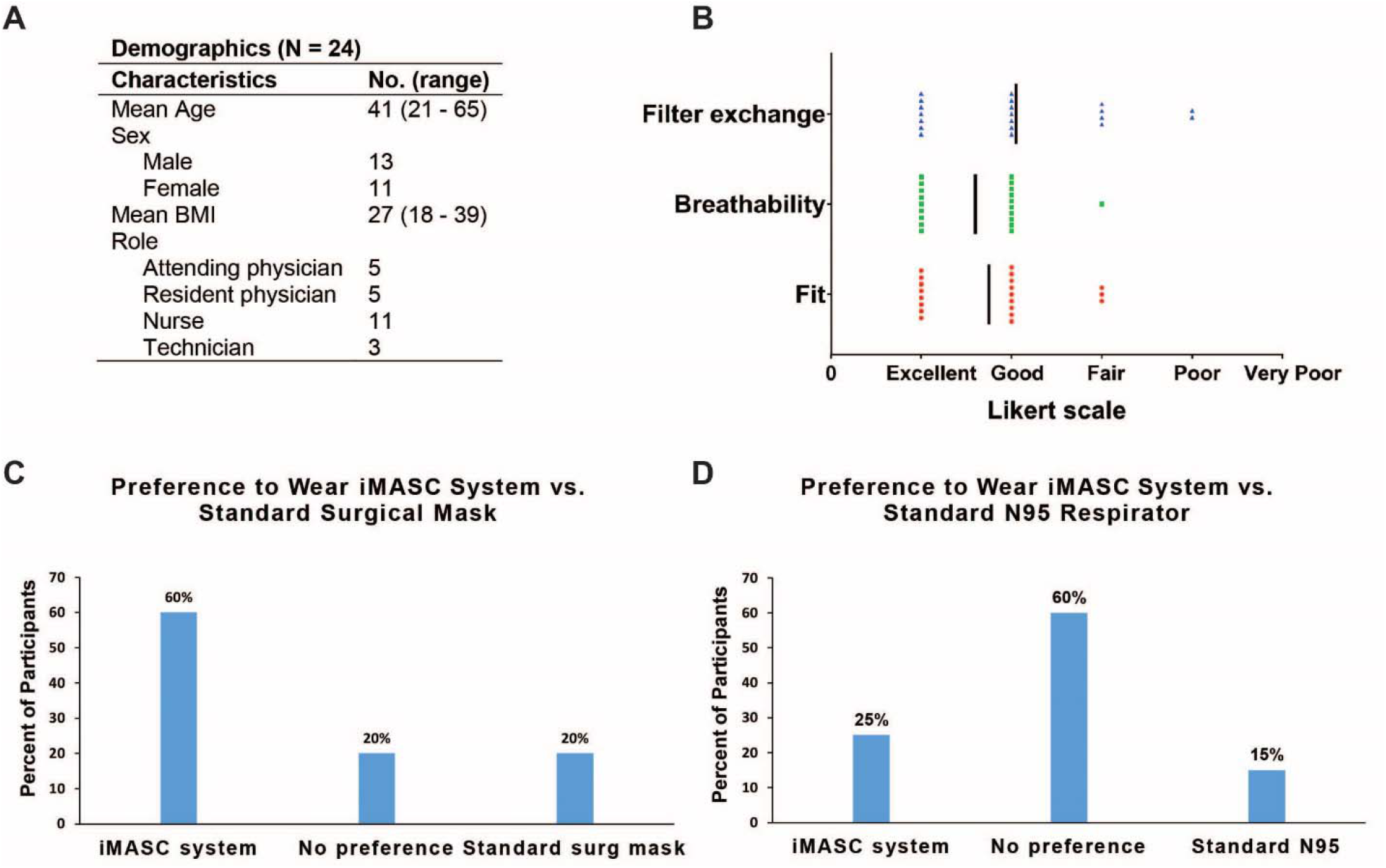
Fit testing of iMASC system in healthcare workers and their user experience. (A) Demographics of participants (N = 24) enrolled in fit testing clinical trial. (B) User experience (N = 20) with the mask based upon a Likert scale. User preferences (N = 20) comparing the iMASC system to the (C) standard surgical mask and (D) N95 respirators.

All participants (n=20) that performed the fit test successfully completed the fit test as part of the hospital annual policy. All participants passed their fit test and were also able to successfully replace the filter into the mask, resulting in a 100% success rate for both fit testing and filter exchange. User experience with the iMASC system was evaluated using a Likert scale with a score of 1 indicating excellent and a score of 5 indicating very poor. Of the 20 participants, the average fit score of the mask was a 1.75 (**Fig. 3B**). Participants on average scored the breathability of the mask as a 1.6 with a median of 1.5. Finally, ease of replacing the filter on the mask was scored on average as a 2.05 with a median score of 2. Participants’ preference to wear the iMASC over a surgical mask or an N95 respirator was also assessed. Sixty percent of participants indicated they would be willing to wear our mask instead of a surgical mask, with 20% indicating no preference between our mask and a standard surgical mask and 20% indicating they would prefer to wear a surgical mask (**Fig. 3C**). When asked about preference to wear our mask instead of an N95 respirator, 25% of participants indicated they would prefer to wear our mask and 60% indicated no preference between our mask and a standard issue mask, with only 15% indicating they would prefer to wear a standard issue N95 respirator (**Fig. 3D**).

## Discussion

During times of pandemics, it is essential to protect healthcare workers from infection and transmission of disease with adequate PPE (*4,9*). As stocks of N95 face masks have reduced, healthcare workers are forced to find alternative strategies of protection, including re-sterilizing masks and using alternative mask materials that may result in less protection and higher disease transmission (*9-10*). Our approach here was to develop a scalable, reusable face mask that can extend the amount of N95 material while providing the same droplet protection as standard N95 masks. The iMASC system was shown to successfully fit multiple different face sizes and shapes using an OSHA approved testing method. Based on the success of the iMASC system in fit testing, this approach could be scaled up for use across many locations. By selecting injection molding as the fabrication technique for the iMASC system, we believe we possess a fundamental advantage to other initiatives using three-dimensional (3D) printing techniques because injection molding is highly scalable and has decreased production time when compared to 3D printing.

These are initial proof-of-concept studies and have some limitations. First, filter replacement was noted to be slightly challenging and additional design changes, such as slight adjustments to dimensions and tolerances, would likely improve the fit and robustness. Additional investigation into user sizing of head straps will be investigated, so as to accommodate more potential users. All post injection-molding manufacturing steps were completed in-house and in large scale production would be outsourced to contracted manufacturers with greater quality control of filter components.

Newer face masks, such as our iMASC system, have potential to resupply hospitals and clinicals with effective N95-comparable masks. Furthermore, a 2018 consensus report from the National Academies of Engineering, Science, and Medicine recommended that the durability and reusability of elastomeric respirators made them desirable for stockpiling for emergencies (*5*). This approach could be applicable to users outside of the healthcare setting, including people in the research, home improvement, and manufacturing settings.

## Materials and Methods

### iMASC fabrication

Masks were designed in SolidWorks based upon current 3M 1860 N95 masks. Once optimized, the design was exported as a SolidWorks file. Reusable face masks were then generated by Protolabs through injection molding out of liquid silicone rubber. Elastic straps were used to secure the mask to the wearer’s face. The mask utilized dual, replaceable filters consisting of virgin N95 filter media bonded to a rigid retaining ring which can easily press-fit into recessed areas of the mask. A 3-inch long, 5mm wide aluminum strip was bonded across the bridge of the nose section of the mask similar to traditional N95 masks.

### Material selection and testing

As a material currently used in anesthesia masks, DOW QP1-250 LSR was selected as a proven injection molding material which enabled greater design freedom for the manufacturing process. Mechanical testing according to ASTM D412 was performed on samples cut directly from masks exposed to a variety of sterilization methods including 10 cycles of autoclaving, 10- minute soak in 10% bleach solution, and 10-minute soak in isopropanol.

### Face scans

To obtain the 3D face geometry of the participants, we developed an IOS application (app) using the TrueDepth camera from an iPhone 11 to capture the face image of the participants. The app employs the ARKit developed by Apple for the use of face tracking in augmented reality to transform a 2D image with depth information into a 3D mesh. The output 3D mesh would then be converted into a solid model for finite element analysis.

### Deformation studies

The commercial FE package ABAQUS/standard 2017 was used for simulating the deformation response of elastic masks. The 3D FE models were constructed by importing the CAD model of the mask from SolidWorks and scanned images of the participant faces. In all the analyses, we discretized the mask using four-node 3D linear tetrahedron elements with hybrid formulation (C3D4H Abaqus element type). The material behavior of the elastomeric mask was captured using an almost incompressible Neo-Hookean hyperelastic model with Poisson’s ratio of v_0 = 0.499 and density of 1.12E3 kg/m3) with directly imported uniaxial test data described in “Material characterization of the medical-grade silicone elastomer “. The scanned faces were imported as 3D Shell Discrete Rigid Element and meshed using three-node 3D rigid triangular elements (R3D3 Abaqus element type). A simplified contact law (“surface to surface” type interaction) was assigned to the model with a penalty friction coefficient 0.2 for tangential behavior and a “hard” contact for normal behavior. The top-middle edge of the mask was positioned to the node at the center of the line connecting the eyes. The “Quasi-static” dynamic implicit solver (*DYNAMIC module in Abaqus) was used. The mask was deformed by applying tensile forces along bands, shown in **fig S3** using SMOOTH step amplitude curve, while completely constraining the motion of the face. The reaction force of the mask against the face as well as contact pressures were recorded as a function of applied load.

### Clinical studies

Institutional Review Board (IRB) approval was obtained prior to any work (Partners IRB 2020P000852). Subjects were comprised of adult Partners Healthcare staff including physicians, residents, nurses, and technicians who were recruited on a voluntary basis. Subjects were enrolled by study staff. Following enrollment and consent, subjects were briefed on the study procedure and then completed a baseline assessment to obtain general demographic information and ensure they had previously been fit tested successfully. Next, two sets of facial measurements were taken: from the tip of the subject’s nose to the base of their chin and across the width of their cheekbones. Each subject’s face was also scanned by a 3D scanner to generate a digital file.

Subjects underwent fit testing in accordance with the protocol outline in the OSHA guidance in Appendix A of 1910.134 using the Gerson Respirator Fit Test kit (part # 065000). In brief, a demonstration was performed to show subjects how to put on a respirator, how it should be positioned on the face, how to set the strap tension and how to determine a proper fit. Subjects then selected a respirator from the two available sizes and adjusted the facepiece until it provided an acceptable fit and was comfortable. Fit was defined as proper placement of the chin; adequate strap tension; fit across the bridge of the nose; tendency of respirator to slip; and ensuring the respirator was of proper size to span between the bridge of the nose and the chin through self-observation in a mirror. Comfort was defined as the position of the mask on the nose, face, and cheeks; room for eye protection; and room to talk. Once the mask was deemed comfortable and of adequate fit, the subject performed a user seal check. To check positive pressure, subjects gently exhaled while wearing the mask to see if the facepiece bulged slightly. Similarly, to perform a negative pressure air check subjects took a deep breath in while wearing the mask and observed for areas of collapse. If air leaked between the subject’s face and the face seal of the respirator or if bulging or collapse occurred during the user seal test, the subject removed the mask and began the procedure again with a new mask. If the subject passed, they proceeded to the fit test.

Subjects first ensured they could detect the taste of the Saccharine test solution. Without a mask on, subjects donned a hood with a fitted collar with a nozzle hole in front of the subject’s mouth and nose. The subject was instructed to breathe through his or her nose and to report when a bitter taste was detected. An inhalation medication nebulizer containing the test solution was gently squeezed ten times while attached to the hood apparatus to aerosolize the test solution into the hood for an approximate volume of 1ml of aerosolized test solution in the hood. If the subject reported a bitter taste, the threshold test was considered complete. If the subject was unable to taste anything, ten more squeezes were administered. Again, if the subject reported a bitter taste the threshold test was considered complete and if not, another ten squeezes were administered (30 total). If the subject was unable to taste the test solution after 30 squeezes, the subject was considered unable to taste the solution and was excused from the study. Study staff recorded the taste threshold indicated in the threshold test for each subject.

After successful completion of the threshold screening test, subjects donned the mask they had previously fitted for comfort and fit under a hood with a fitted collar and were instructed to report if they could taste the test solution. A nebulizer of odorous solution (Saccharin) was inserted into the hole in the front of the hood and sprayed at the same concentration (10, 20, or 30 squeezes) as the subject was able to taste in their initial threshold test. The subject was instructed to perform the following exercises while the aerosolized solution was replenished every 30 seconds: normal breathing, deep breathing, turning the head side to side, moving the head up and down, counting backwards from 100, grimacing, bending over, and finally normal breathing for a second time. If the subject at any time during the fit test was able to taste the solution, they indicated to the study staff and the test was considered failed. If the subject did not report tasting the solution the test was considered passed.

Subjects who passed the fit test were introduced to how to properly replace the filter with a demonstration by study staff. Subjects were then asked to replace the filter and perform a user seal check to ensure an adequate fit. Subjects then performed a second fit test with the replacement filter. Finally, subjects completed an exit assessment where they ranked fit, breathability, and difficulty of replacing the filter according to a Likert scale. Subjects were also asked about their willingness to wear the mask compared to either a surgical mask and an N95 mask. All testing was performed at Brigham and Women’s Hospital.

## Data Availability

All data referred to in the manuscript is available in the manuscript and the supplementary materials.

https://imasc.mit.edu

## Supplementary Materials

**Fig. S1.**
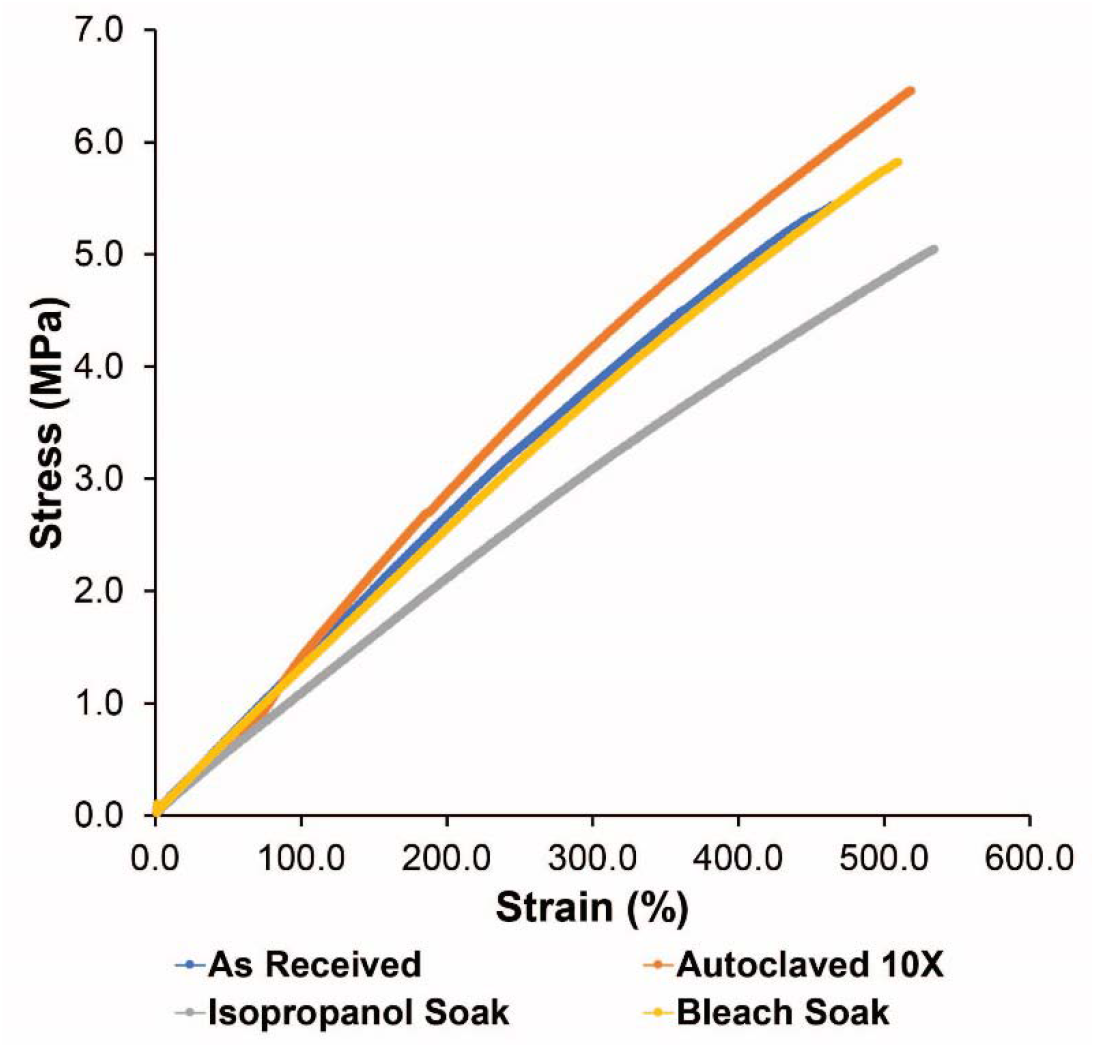
Mechanical testing on samples cut directly from masks exposed to a variety of sterilization methods including 10 cycles of autoclaving, 10-minute soak in 10% bleach solution, and 10-minute soak in isopropanol.

**Fig. S2.**
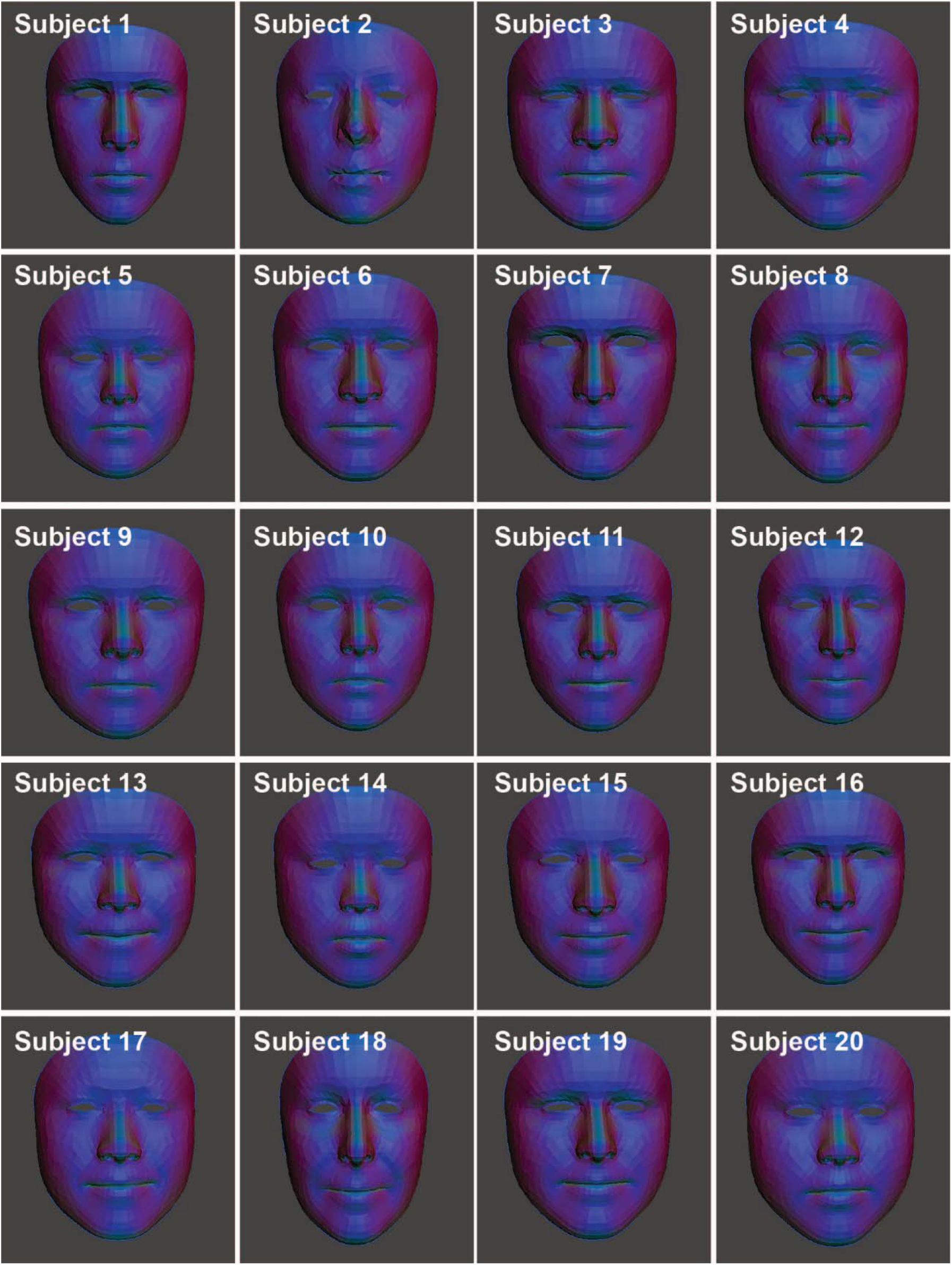
Front view of 3D facial reconstruction of participants faces in fit trial of the iMASC system.

**Fig. S3.**
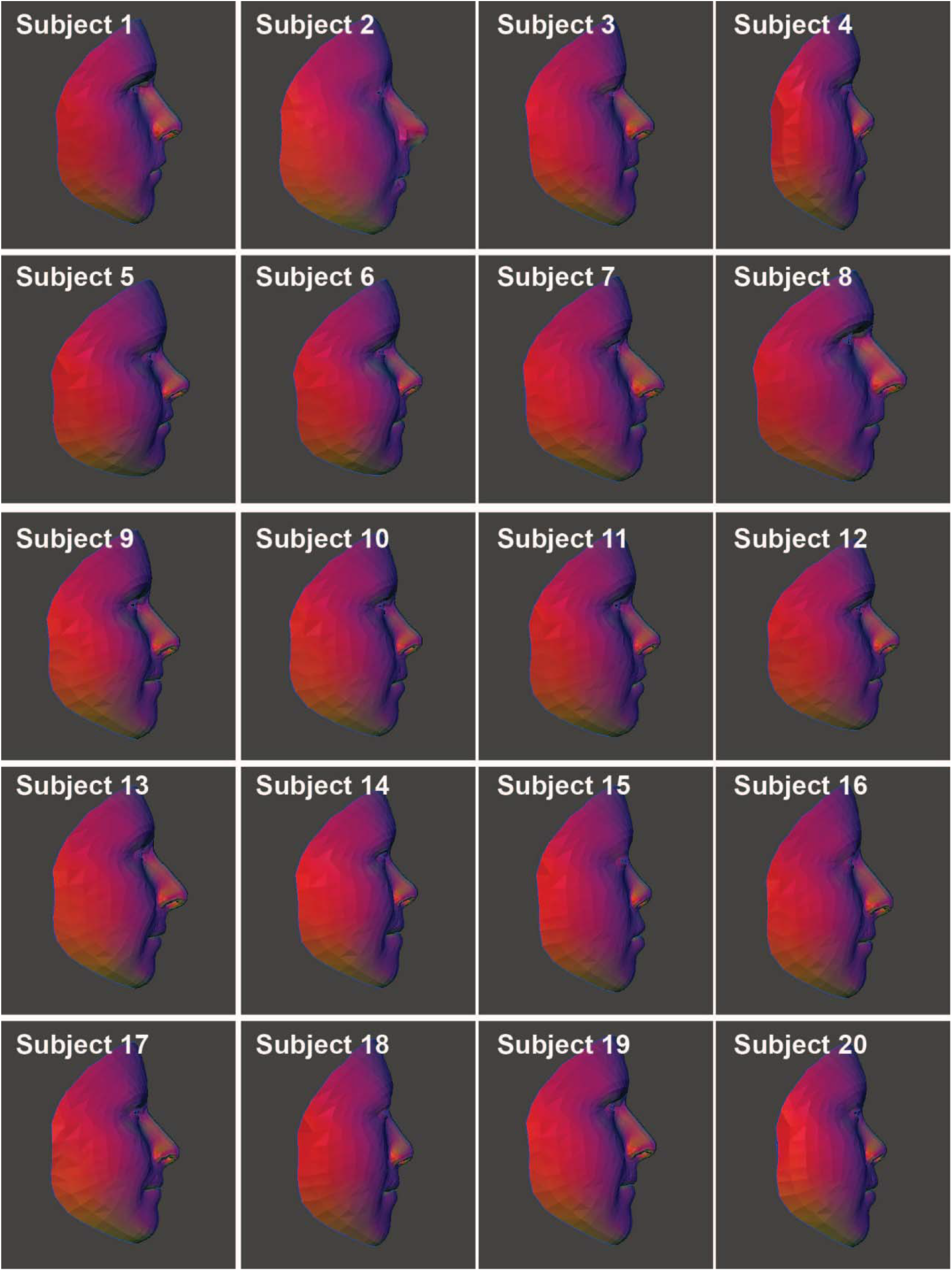
Side view of 3D facial reconstruction of participants faces in fit trial of the iMASC system.

**Fig. S4.**
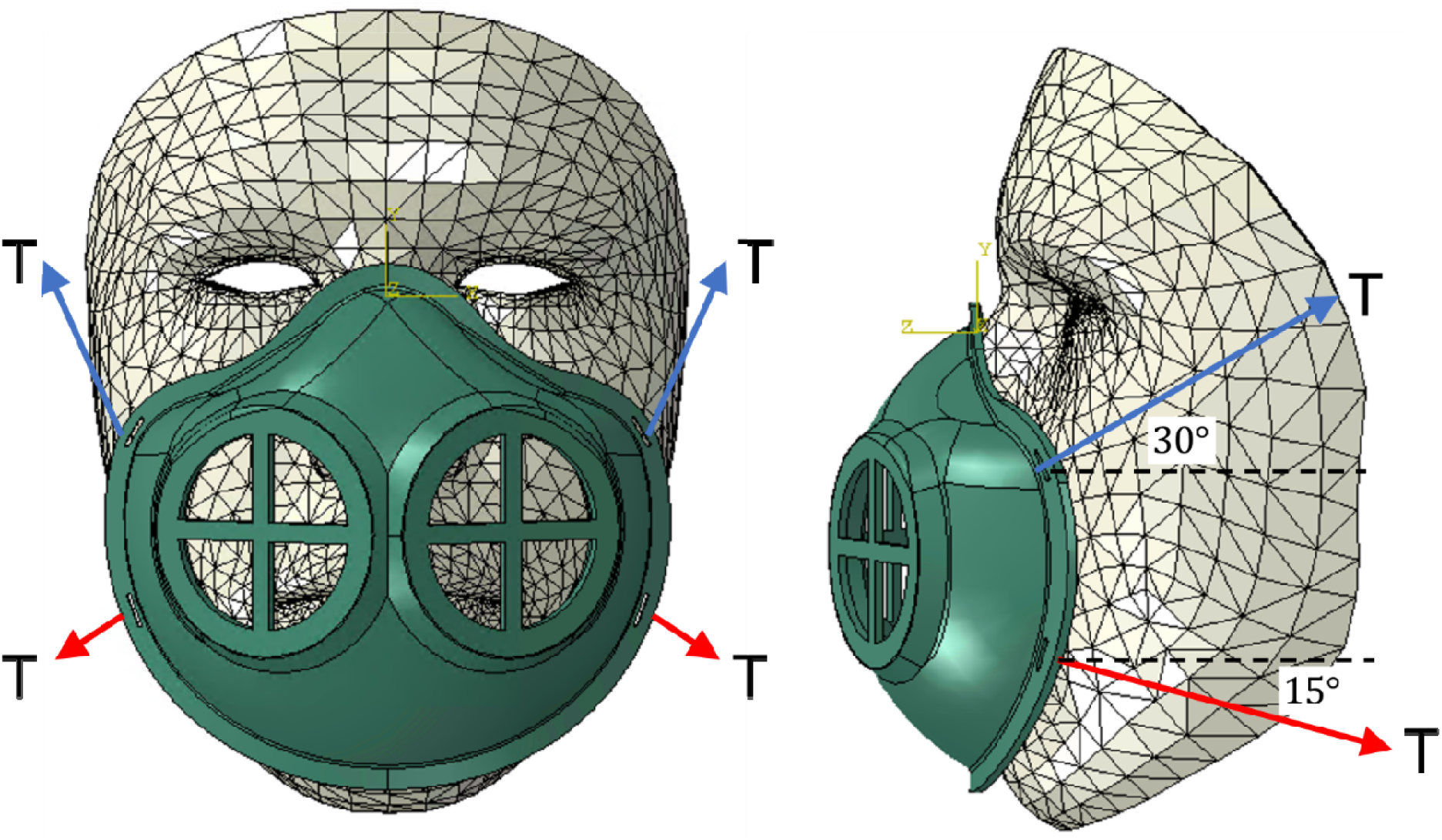
Illustration of the applied loads via mask straps.

**Table S1.**
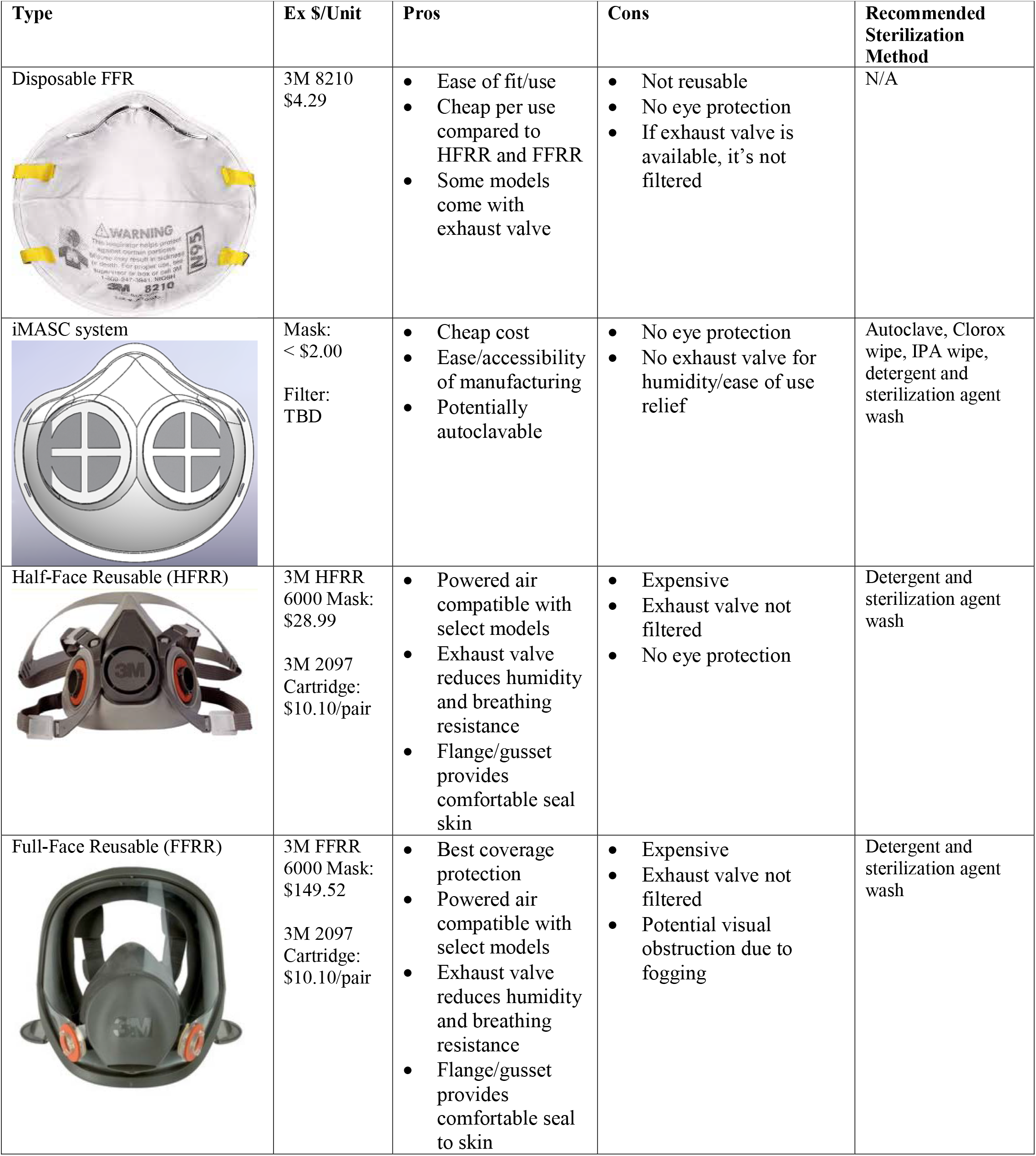
Array of N95 and N95-comparable technologies.

## Acknowledgments

We thank Ania Hupalowska for her illustrations of the clinical workflow. We thank Prof. R. Langer for helpful discussions around mask development.

## Contributions

J.D.B. and A.J.W. designed and fabricated the iMASC system, assisted with the clinical trial, analyzed and interpreted data, and wrote the manuscript. P.R.C. performed the clinical trial, analyzed and interpreted data, and wrote the manuscript. H.W.H. and S.B. designed the face scanning and performed FEA modeling, analyzed data, and wrote the manuscript. S.B., C.T., and S.M. analyzed data and designed prototypes. G.T. supervised, reviewed the data and edited the manuscript.

## Data Availability

The authors declare that the data supporting the findings of this study are available within the paper and its supplementary information files.

## References and Notes

1. M. L. Ranney, V. Griffeth, A. K. Jha. Critical Supply Shortages - The Need for Ventilators and Personal Protective Equipment during the Covid-19 Pandemic. N. Engl. J. Med. 2020. doi: 10.1056/NEJMp2006141.

2. E. Livingston, A. Desai, M. Berkwits. Sourcing Personal Protective Equipment During the COVID-19 Pandemic. JAMA. (2020) doi: 10.1001/jama.2020.5317.

3. J. G. Adams, R. M. Walls. Supporting the Health Care Workforce During the COVID-19 Global Epidemic. JAMA. (2020) doi: 10.1001/jama.2020.3972.

4. The Lancet. COVID-19: protecting health-care workers. Lancet. 395, 922 (2020).

5. National Academies of Sciences, Engineering, and Medicine; Health and Medicine Division; Board on Health Sciences Policy; Committee on the Use of Elastomeric Respirators in Health Care; Reusable Elastomeric Respirators in Health Care: Considerations for Routine and Surge Use. Liverman CT, Yost OC, Rogers BME, et al., editors. Washington (DC): National Academies Press (US); 2018.

6. A. K. Brill, R. Pickersgill, M. Moghal, M. J. Morrell, A. K. Simonds. Mask pressure effects on the nasal bridge during short-term noninvasive ventilation. ERJ Open Res. 4, 00168–2017 (2018).

7. Occupational Safety and Health Standards. Appendix A to §1910.134—Fit Testing Procedures (Mandatory).

8. Temporary Enforcement Guidance □ Healthcare Respiratory Protection Annual Fit□Testing for N95 Filtering Facepieces During the COVID□19 Outbreak. 2020.

9. S. Feng, C. Shen, N. Xia, W. Song, M. Fan, B. J. Cowling. Rationale use of face masks in the COVID-19 pandemic. Lancet Respir. Med. (2020) doi: 10.1016/S2213-2600(20)30134-X.

10. C. R. MacIntyre, H. Seale, T. C. Dung, N. T. Hien, P. T. Nga, A. A. Chughtai, B. Rahman, D. E. Dwyer, Q. Wang. A cluster randomised trial of cloth masks compared with medical masks in healthcare workers. BMJ Open. 5, e006577 (2015).

